# Effects of α-synuclein pathology in normal aging and Alzheimer’s disease

**DOI:** 10.1101/2025.08.06.25333152

**Authors:** Joseph R. Winer, Melanie J. Plastini, America Romero, Hillary Vossler, Isha Sai, Divya Channappa, Carla Abdelnour, Marian Shahid-Besanti, Edward N. Wilson, Christina B. Young, Alexandra Trelle, Maya Yutsis, Sharon Sha, Veronica Ramirez, Ryan Taylor, Kyan Younes, Tony Wyss-Coray, Victor W. Henderson, Anthony D. Wagner, Kathleen L. Poston, Elizabeth C. Mormino

## Abstract

**Objective:** α-synuclein is the hallmark pathology of Parkinson’s disease and dementia with Lewy bodies, described together as Lewy body disease (LBD). α-synuclein is also commonly observed in the context of Alzheimer’s disease (AD). Here we investigate the frequency of α-synuclein biomarker positivity in clinically unimpaired (CU) individuals and an AD research cohort, as well as associations with demographics, AD biomarkers, cognitive performance, and clinical outcomes.

**Methods:** We assessed α-synuclein status (α-syn+/-) in 270 CU and 56 clinically diagnosed AD participants (29 mild cognitive impairment and 27 dementia) using a cerebrospinal fluid seed amplification assay (α-syn SAA). Eighty-five LBD spectrum participants were included for comparison. AD biomarker levels were measured with cerebrospinal fluid β-amyloid42/40 and p-tau181. Participants received cognitive testing, the Neuropsychiatric Inventory Questionnaire and the Movement Disorders Society Unified Parkinson’s Disease Rating Scale.

**Results:** α-syn was detected in 9% of CU, 14% of AD mild cognitive impairment, and 19% of AD dementia participants, whereas 81% of individuals with LBD spectrum clinical diagnoses were α-syn+. α-syn+ CU were older, performed worse on tests of executive function and working memory, and reported more LBD-related non-motor symptoms relative to α-syn-CU. α-syn status in CU was not significantly associated with β-amyloid or tau, memory performance, motor symptoms, or neuropsychiatric symptoms.

**Conclusions:** Using the CSF SAA biomarker, α-syn positivity independently predicts subtle cognitive changes and early clinical symptoms in aging. These cross-sectional findings represent an important addition to the limited but growing literature characterizing the frequency and effects of α-syn positivity in clinically healthy older adults and individuals with AD.

## Introduction

Parkinson’s disease (PD) and dementia with Lewy bodies (DLB) are neurodegenerative syndromes collectively referred to as Lewy body diseases (LBD) that are pathologically characterized by the intraneuronal aggregation of misfolded α-synuclein. In addition to motor and cognitive syndromes, individuals with α-synuclein pathology can develop “prodromal” symptoms such as isolated REM-sleep behavior disorder and other non-specific non-motor symptoms^1^. Critically, α-synuclein pathology also occurs in the context of other neurodegenerative syndromes, such as Alzheimer’s disease (AD)^2^. Given the broad clinical presentations seen in synucleinopathies, accurate biomarkers are crucial to understand the effects of underlying α-synuclein pathophysiology across the clinical spectrum^3^.

Fortunately, recent developments allow for robust detection of α-synuclein pathology *in vivo* through cerebrospinal fluid (CSF) α-synuclein seed amplification assay (α-syn SAA)^4^ or skin biopsy^5^. The α-syn SAA is exquisitely sensitive, capable of detecting a single molecule of misfolded α-synuclein, and shows high accuracy in detecting cortical Lewy body deposition in the context of LBD spectrum diagnoses compared to postmortem neuropathology^6–8^. Such advances in highly-sensitive biomarkers have motivated calls for biologically defining all individuals found to have neuronal synuclein disease (NSD), anchored to markers of α-synuclein aggregation and dopaminergic dysfunction, and staging according to clinical symptoms and functional impairment^3^. One opportunity inherent to this proposed staging framework is the concept of a biomarker-defined “preclinical” (or presymptomatic) stage of NSD. A precedent for this framework comes from the AD field, which conceptualizes clinically unimpaired (CU) individuals with biomarker evidence of abnormal β-amyloid (Aβ) as a preclinical stage of AD^9^. To date, studies leveraging CSF α-syn SAA data suggest an α-syn positivity prevalence between 8-16% in CU older individuals^10–12^, which is similar to estimates from postmortem studies on the presence of Lewy bodies anywhere in the brain^13–15^. Subtle effects related to cognition as well as other non-cognitive LBD features have been explored in an emerging literature examining CU α-syn+ individuals^10–12,16^. However, reported effects and measures are not consistent across studies, highlighting the need for additional data characterizing the early effects of α-syn positivity.

We investigated the frequency of CSF α-syn SAA positivity in a cohort of CU older adults, individuals with AD, and individuals with clinical LBD. We also examined associations between α-syn positivity and demographics, AD biomarkers, cognitive test performance, and clinical measures. We hypothesized that α-syn+ CU older adults would perform worse in cognitive domains typically associated with LBD, such as executive function. We further hypothesized that α-syn+ CU individuals would be more likely to have elevated AD biomarkers as well as non-motor and motor symptoms related to LBD.

## Methods

### Participants

We studied N=416 participants from studies at Stanford University that collected CSF between 2010 and 2023, including the Stanford Alzheimer’s Disease Research Center (ADRC), the Stanford Aging and Memory Study (SAMS), and the Pacific Udall Center (PUC). All participants underwent lumbar puncture and had α-syn SAA data available.

All participants underwent diagnostic adjudication at multidisciplinary consensus meetings, which included a panel of neurologists, neuropsychologists, and research staff. All CU participants were older adults who did not meet criteria for PD, mild cognitive impairment, or dementia based on history and neurological examination findings. All participants diagnosed clinically with mild cognitive impairment due to AD (AD-MCI) or dementia due to AD (AD-Dementia) met criteria for National Institutes of Health Alzheimer’s Disease Diagnostic Guidelines^17,18^ and were confirmed to have biomarker evidence of AD using CSF Aβ42/40 (see criteria below). Ten participants with AD diagnoses were removed from analyses, 3 who were CSF Aβ42/40 negative and 7 who had no CSF Aβ42/40 data available. PD was diagnosed using the UK Brain Bank criteria and required bradykinesia with muscle rigidity and/or rest tremor^19^. Participants were further categorized as having PD with mild cognitive impairment (PD-MCI) if they had a cognitive complaint and objective impairment on comprehensive cognitive testing using MDS Level II criteria^20^ without substantial impact on functional activities. PD dementia (PDD) was defined as cognitive impairment severe enough to interfere with activities of daily living^21^ as determined by clinical history and the Clinical Dementia Rating^22^. Mild cognitive impairment due to Lewy bodies (MCI-LB) and DLB were defined according to published criteria^23,24^. Study participants were grouped into three clinical etiological categories: CU, AD (AD-MCI and AD-Dementia), and LBD (PD without cognitive impairment, PD-MCI, MCI-LB, PDD, and DLB). Five ADRC participants with α-syn SAA results did not meet clinical criteria for CU, AD, or LBD. These participants were not included in analyses and are described in **Supplementary Table 1**. The Stanford Institutional Review Board approved this study, and all study participants provided written informed consent.

### CSF collection and assessment of Aβ and tau

CSF collection procedures for ADRC, PUC, and SAMS followed a standard operating procedure so that all samples were collected, banked, and stored in a similar fashion. CSF was collected into polypropylene tubes by lumbar puncture before 11 AM. CSF was stored in 1.0 or 0.5 mL aliquots at −80°C until analysis. The fully automated Lumipulse *G* 1200 instrument (Fujirebio US, Inc., Malvern, PA) was used to measure CSF p-tau181, Aβ42, and Aβ40 as described previously^25,26^. In a dataset of 291 participants with a diagnosis of CU, AD-MCI, or AD-Dementia, a 2-cluster Gaussian mixture modeling approach was used to define a Aβ42/Aβ40 cutoff of 0.0943 (based off the 0.5 probability of belonging to the Aβ+ distribution)^25^. The p-tau181 cutoff of 57.5 pg/mL was determined as 2 standard deviations above the mean of Aβ-CU in the current dataset. Aβ42/Aβ40 data were unavailable for 19 participants (16 CU, 3 LBD) and p-tau181 data were unavailable for the same 19 as well as an additional 2 CU participants; these participants were excluded from all analyses that included AD biomarkers.

### α-synuclein seed amplification assay

CSF samples were analyzed in Amprion Inc.’s CLIA-certified (05D2209417) and CAP-accredited (8168002) laboratory (San Diego, CA) in two batches that utilized the same methodology. See **Supplementary Methods** for an overview of the samples comprising the two batches. Clinical laboratory personnel who performed the α-syn SAA were blinded to all clinical diagnoses and demographic data associated with the samples. Participants’ samples were analyzed using the qualitative version of α-syn SAA that has been validated for clinical use under CAP/CLIA (SYNTap Biomarker Test) for the presence of misfolded α-synuclein aggregates as previously described^27,28^. Briefly, each sample was run in triplicate using 40 μl CSF per well in a COSTAR 96 well plate with a final reaction volume of 200 μl. Each reaction mixture contained 0.3 mg/mL recombinant α-synuclein in 100 mM PIPES pH 6.50, 500 mM NaCl, 10 mM ThT, and a 2.5 mm borosilicate glass bead per well. A baseline fluorescence reading was taken using a BMG LABTECH FLUOStar Ω Microplate Reader. Plates were then transferred to a TIMIX 5 shaker placed in an incubator set to 37°C and subjected to cycles of orbital shaking at 800 rpm for 1 min followed by 29 min of quiescent incubation for 7-10 days. Fluorescence readings were repeated once per day throughout the reaction period. Following final measurement, the maximum relative fluorescence units of each well was determined and the median of the three wells for each sample was calculated. CSF samples were classified as “detected” or “not detected” based on a preestablished threshold for the median maximum fluorescence of the triplicate wells. The highest raw fluorescence from each well was used in a probabilistic algorithm to establish whether each of the three replicates was a positive or negative readout. There were three possible outcomes: 1) if all three replicates from a sample were detected, the sample was classified as α-synuclein positive (α-syn+); 2) if zero or one of the replicates were detected, the sample was classified as α-synuclein negative (α-syn-), 3) if two replicates were positive, the sample was deemed indeterminate. As a secondary classification criterion, samples with inconclusive results, defined by high replicate variability or low average maximum ThT fluorescence across the three replicates, were re-evaluated and those falling below the established fluorescence threshold were classified as negative.

Samples from two CU participants were classified by Amprion, Inc. as detected during an extended assay window beyond current validated specifications. We designated these two participants as α-syn+ in all analyses.

### Genetics data

Apolipoprotein E (APOE) genotyping was determined from whole-genome sequencing (WGS) or obtained from National Cell Repository for Alzheimer’s Disease (NCRAD) using a Fluidigm fingerprint panel. WGS was performed at the Beijing Genomics Institute in Shenzhen, China, or sequenced as part of the Stanford Extreme Phenotypes in AD project with sequencing performed at the Uniformed Services University of the Health Sciences (USUHS) on an Illumina HiSeq platform. The Genome Analysis Toolkit (GATK) workflow Germline short variant discovery was used to map genome sequencing data to the reference genome (GRCh38) and to produce high-confidence variant calls using joint-calling^29^. APOE genotype (ε2/ε3/ε4) was determined using allelic combinations of single nucleotide variants rs7412 and rs429358. APOE genotype was available for 340 participants (218 CU, 44 AD, and 78 LBD).

### Cognitive and clinical assessments

Since neuropsychological batteries differed between study designs, analyses focused on cognitive tests common to CUs across all cohorts: Digit Span forward and backward (N=241), Trail Making Test (Trails A duration, N=263; and Trails B-A, N=262), Hopkins Verbal Learning Test - Revised (HVLT-R) Delayed Recall (N=241), and semantic fluency (N=265). Digit Span data were comprised of Uniform Data Set Number Span in Stanford ADRC and PUC and WMS-III Digit Span in SAMS, so WMS-III scores were capped at the maximum Number Span scores of 14 for forward and 12 for backward. Cognitive analyses were restricted to CU due to insufficient data being available from MCI-AD and Dementia-AD participants.

Non-motor and motor symptom severity were evaluated in a subset of participants using the Movement Disorder Society-sponsored revision of the Unified Parkinson’s Disease Rating Scale (MDS-UPDRS). MDS-UPDRS scores were available from N=83 (N=7 α-syn+) CU participants and N=20 (N=2 α-syn+) participants with AD for Part I (non-motor experiences of daily living), N=82 (N=6 α-syn+) CU and N=19 (N=2 α-syn+) AD for Part II (motor experiences of daily living), and N=96 (N=8 α-syn+) CU and N=18 (N=2 α-syn+) AD for Part III (motor examination). Neuropsychiatric symptoms were evaluated in a subset of participants using the Neuropsychiatric Inventory-Questionnaire (NPI-Q; N=195 [N=19 α-syn+] CU, N=49 [N=7 α-syn+] AD). Item severity scores were summed to create a total score representing overall neuropsychiatric symptom severity.

### Statistical analysis

We performed logistic regression analyses to determine associations between demographic variables, AD biomarkers, and α-syn SAA status. Separate models examined these associations within CU alone, AD (AD-MCI and AD-Dementia) alone, and CU and AD together. Models with the AD group included cognitive status as an additional covariate (CU, AD-MCI, AD-Dementia). Logistic regression was also used to determine odds ratios for α-syn positivity predicting Aβ and tau positivity in CU, and for APOE ε4 carriage predicting α-syn positivity in CU, all adjusting for age. To investigate the relationship between α-syn positivity and cognition in CU, we performed linear regression analyses with cognitive test scores as the dependent variable and α-synuclein status, age, sex, years of education, and CSF p-tau181 as independent variables. For clinical assessments, MDS-UPDRS scores and NPI-Q total scores were dependent variables in linear regression models with α-syn SAA status, age, and sex as independent variables. For MDS-UPDRS and NPI-Q, separate models examined these associations within CU alone and CU and AD together, with models with the AD group again including a term for cognitive status. We considered 2-sided *p* < .05 to be statistically significant. No multiple comparisons correction was performed. All statistical analyses were performed in R version 4.4.1.

## Results

### Participants and frequency of α-synuclein pathology

Participant demographics for CU (N=270) and AD (N=56) groups are summarized in the **Table**. Frequency of α-syn positivity is visualized together with the frequency of Aβ and tau positivity across all diagnostic groups in **Figure 1**. CSF α-syn SAA determined that 24 (8.9%) CU, 4 (13.8%) AD-MCI, and 5 (18.5%) AD-Dementia participants were α-syn+. Within the clinical LBD groups (N=85; **Supplementary Table 2**), 69 (81.1%) were α-syn+. 88 (35%) CU and 35 (43%) LBD participants were Aβ+ by CSF Aβ42/40 ratio, and 43 (17%) CU, 20 (69%) AD-MCI, 25 (93%) AD-Dementia, and 18 (22%) of LBD participants were tau+ by CSF p-tau181.

**Figure 1.**
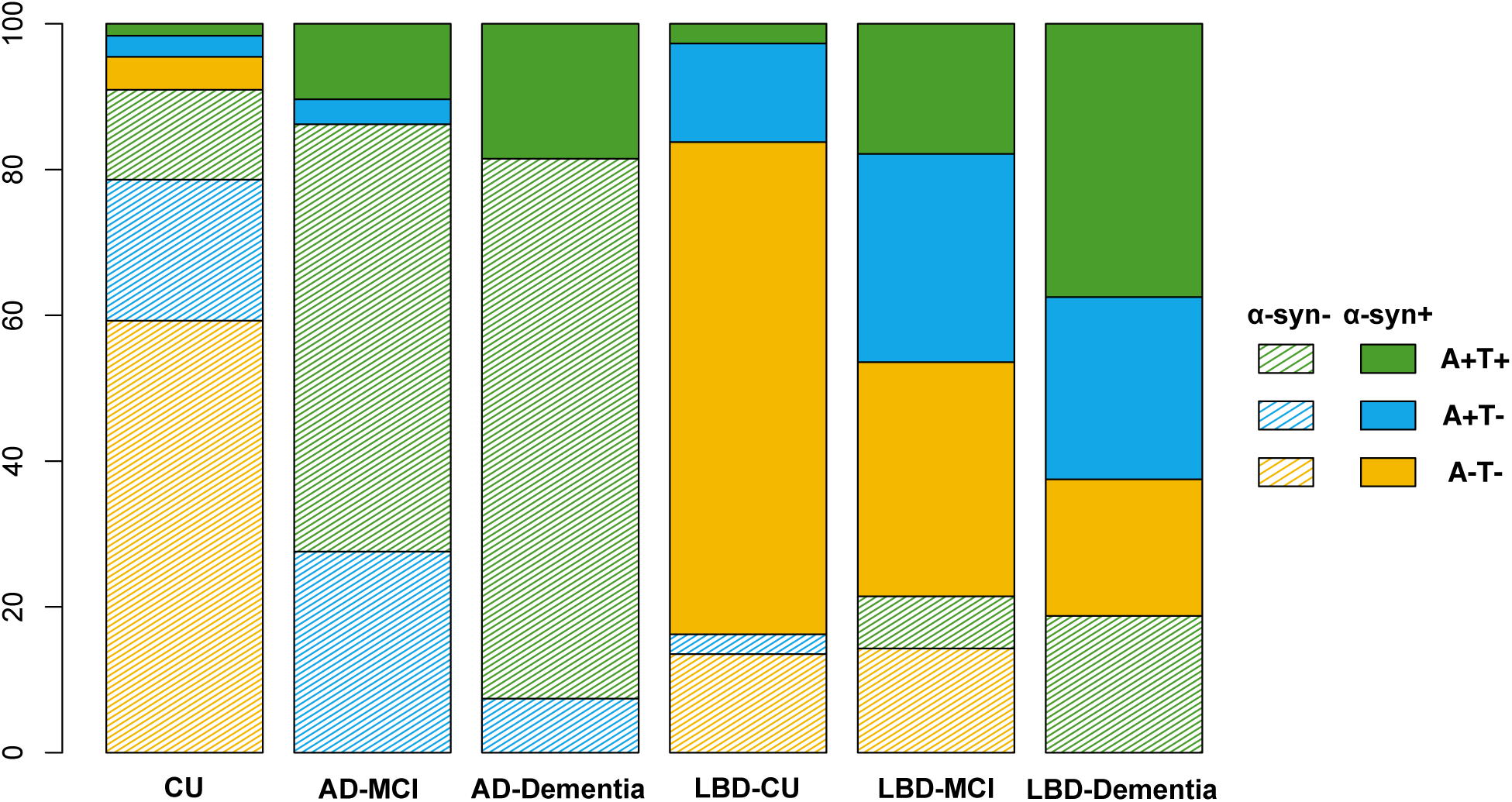
Frequencies of α-synuclein, β-amyloid, and tau positivity across cognitively unimpaired participants and individuals diagnosed with Alzheimer’s disease and Lewy body disease. Bar plot shows percentage for α-synuclein, β-amyloid, and tau status within each clinical group, by etiology and impairment status. A-T+ individuals (N=9) are not represented.

### Associations with AD pathology, demographics, and APOE genotype

First, we investigated associations with α-syn SAA status in CU participants using a logistic regression model that included age, sex, years of education, CSF Aβ42/40 ratio, and CSF p-tau181 as predictors. In this model only older age was a significant predictor of α-syn SAA status (odds ratio [OR] 1.78, 95% CI 1.10-2.97; **Figure 2A**). Next, in a model restricted to participants with AD, that included MCI versus dementia status as an additional predictor, none of the variables were significant in predicting α-syn SAA status (**Figure 2B**). In a third model that included all CU, AD-MCI, and AD-Dementia participants, none of the variables were significant predictors of α-syn SAA status, but older age was marginally significant (OR 1.51, 95% confidence interval [CI] 1.00-2.33; **Figure 2C**).

**Figure 2.**
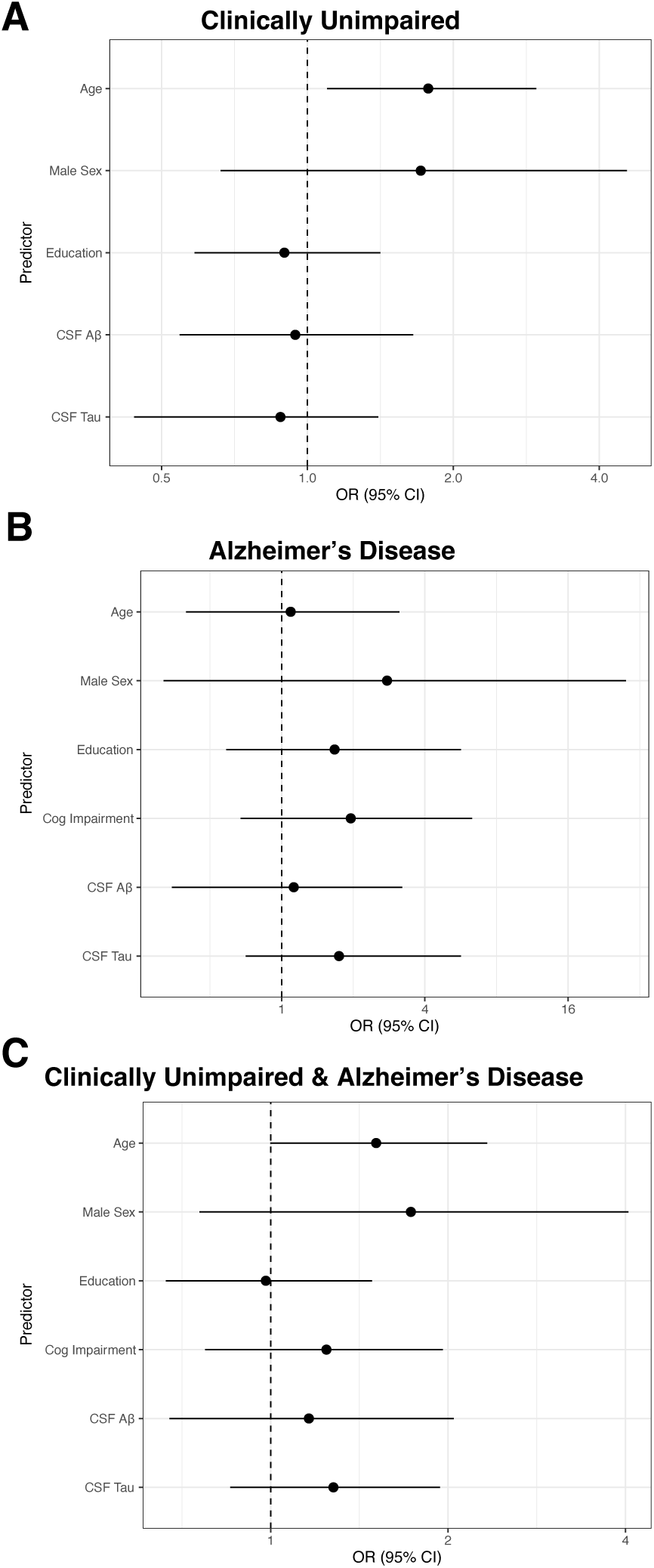
α-synuclein status associations with demographics, β-amyloid, and tau in clinically unimpaired participants and Alzheimer’s disease. Forest plots show adjusted odds ratios (OR) and 95% confidence intervals (CI) predicting α-synuclein positivity for (A) clinically unimpaired participants, (B) participants with Alzheimer’s disease, and (C) both groups together from logistic regression models. Abbreviations: CSF Aβ, CSF β-amyloid 42/40 ratio.

Logistic regressions with dichotomous Aβ and tau status as predictors showed that neither Aβ positivity nor tau positivity, adjusted for age, was significantly associated with α-syn SAA status in CU (Aβ, OR=1.58, 95% CI 0.65-3.82; tau, OR=1.05, 95% CI 0.32-2.93). We additionally examined whether continuous CSF Aβ and tau differed by α-syn SAA status in the CU group. Linear regressions that included α-syn SAA status, age, and sex as independent variables showed no significant association with α-syn SAA status for either AD biomarker (Aβ42/40 ratio, unstandardized β ± standard error = -0.05 ± -0.22, p=0.98; p-tau181, -0.10 ± -0.21, p=0.63; **Figure 3**).

**Figure 3.**
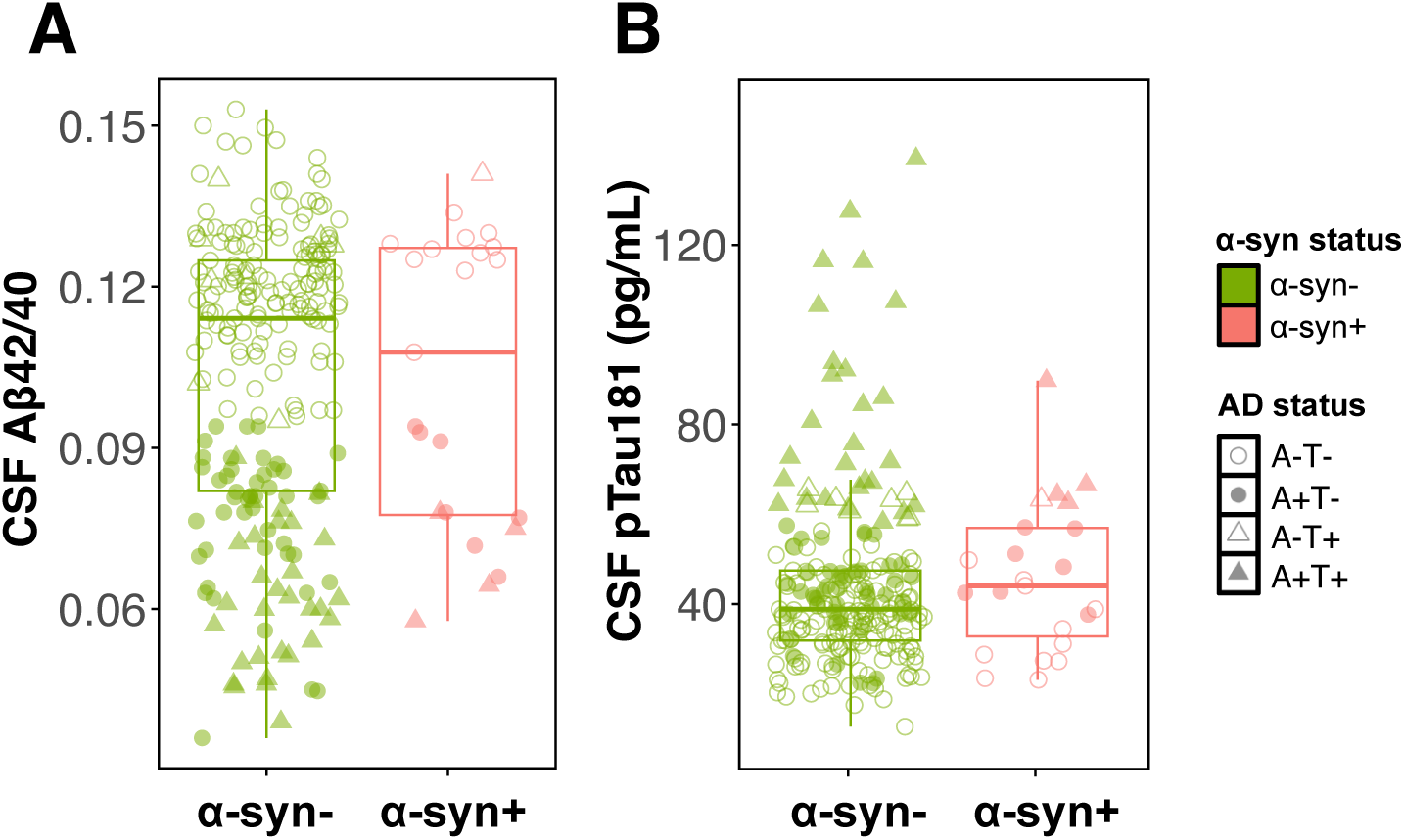
Associations between α-synuclein status and continuous β-amyloid and tau in clinically unimpaired participants. Boxplots show median and interquartile range in (A) CSF β-amyloid 42/40 ratio and (B) CSF p-tau181 for α-synuclein positive and negative clinically unimpaired participants. Unadjusted data are displayed. Group differences are not statistically significant in a linear regression adjusting for age and sex. CSF p-tau181 values for three α-synuclein negative outliers (>165 pg/mL) are omitted from the plot.

Finally, in CU, APOE ε4 carriers were more than twice as likely to be α-syn+, which was marginally significant adjusting for age (OR=2.48 95% CI 0.87-6.56). This effect was attenuated when including Aβ status in the model (APOE ε4, OR=1.92, 95% CI 0.65-5.61, Aβ status OR=1.67, 95% CI 0.57-4.92). Out of N=21 APOE ε4/ε4 homozygotes in the full study sample, 10 (48%) were α-syn+.

### Associations with cognition

We examined associations between cognition and α-syn SAA status within the CU group. Models included α-syn SAA status, age, sex, years of education, and CSF p-tau181. Test performance by α-syn SAA status and linear regression results are displayed in **Supplementary Table 3**. α-syn positivity was associated with worse performance on Trails B-A, a test of executive function (0.44 ± 0.19, p=0.02; **Figure 4A**), and marginally negatively associated with Digit Span Backward, a test of working memory (-0.43 ± 0.23, p=0.07; **Figure 4B**). P-tau181, but not α-syn positivity, was associated with HVLT-R Delayed Recall (-0.21 ± 0.07, p=0.001; **Figure 4C**) and Trails A (0.15 ± 0.07, p=0.04; **Figure 4D**). Semantic fluency and Digit Span Forward performance were not associated with either α-syn SAA status or p-tau181.

**Figure 4.**
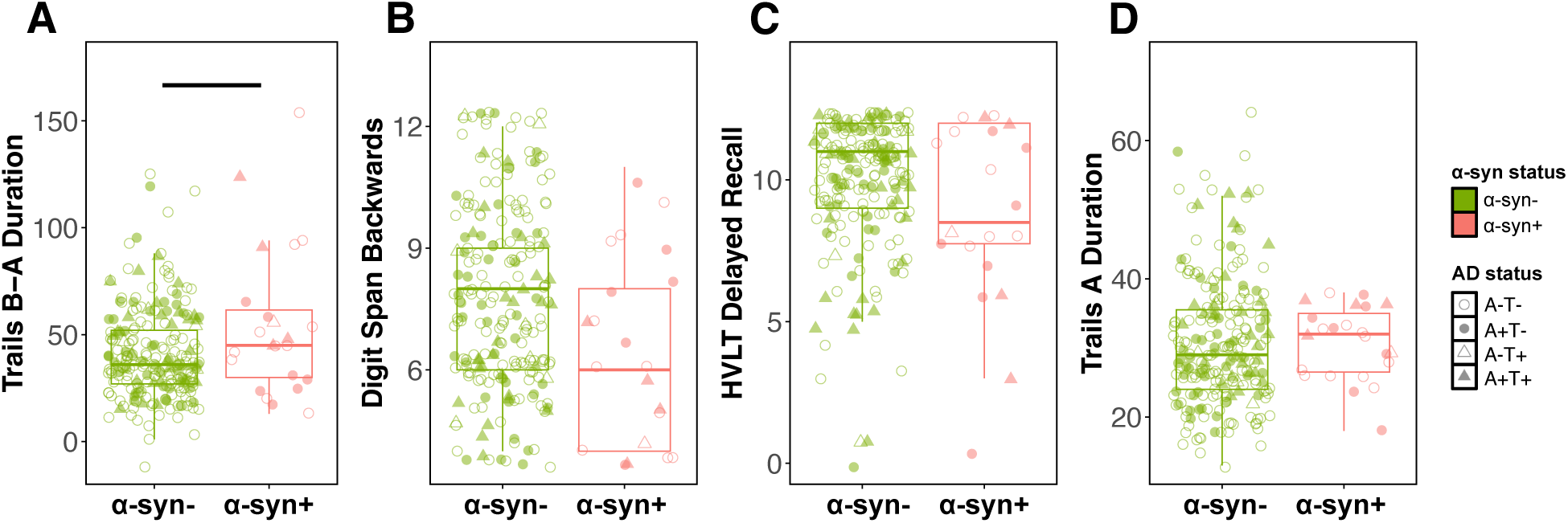
α-synuclein associations with cognitive test performance in clinically unimpaired participants. Boxplots show median and interquartile range in (A) Trail Making Test B-A, (B) Digit Span Backwards, (C) HVLT-R Delayed Recall, and (D) Trail Making Test A performance for α-synuclein positive and negative clinically unimpaired participants. Unadjusted data are displayed. Solid line indicates significant difference in a linear model adjusting for age, sex, years of education, and CSF p-tau181.

### Associations with clinical outcomes

We next examined associations between CSF a-syn status and LBD-related clinical outcomes assessed using the MDS-UPDRS and neurologist-rated motor scores. In a subset of CU with MDS-UPDRS data available, a linear regression adjusting for age and sex revealed that α-syn positivity was associated with higher severity in non-motor symptoms, as assessed by MDS-UPDRS Part I, which includes mood, sleep, and constipation (0.97 ± 0.38, p=0.01, **Figure 5**). Motor symptom scores (self-rated MDS-UPDRS Part II and neurologist-rated MDS-UPDRS Part III) were not significantly associated with α-syn SAA status in CU (Part II, 0.61 ± 0.41, p=0.14; Part III, 0.29 ± 0.36, p=0.43). When individuals with AD-MCI and AD-Dementia were included in the analysis (with diagnosis included as a covariate in the model), α-syn positivity was again significantly associated with non-motor symptom scores (1.04 ± 0.31, p=0.001) and self-rated motor scores (0.87 ± 0.29, p=0.003), but not significantly associated with neurologist-rated motor scores (0.52 ± 0.32, p=0.11).

**Figure 5.**
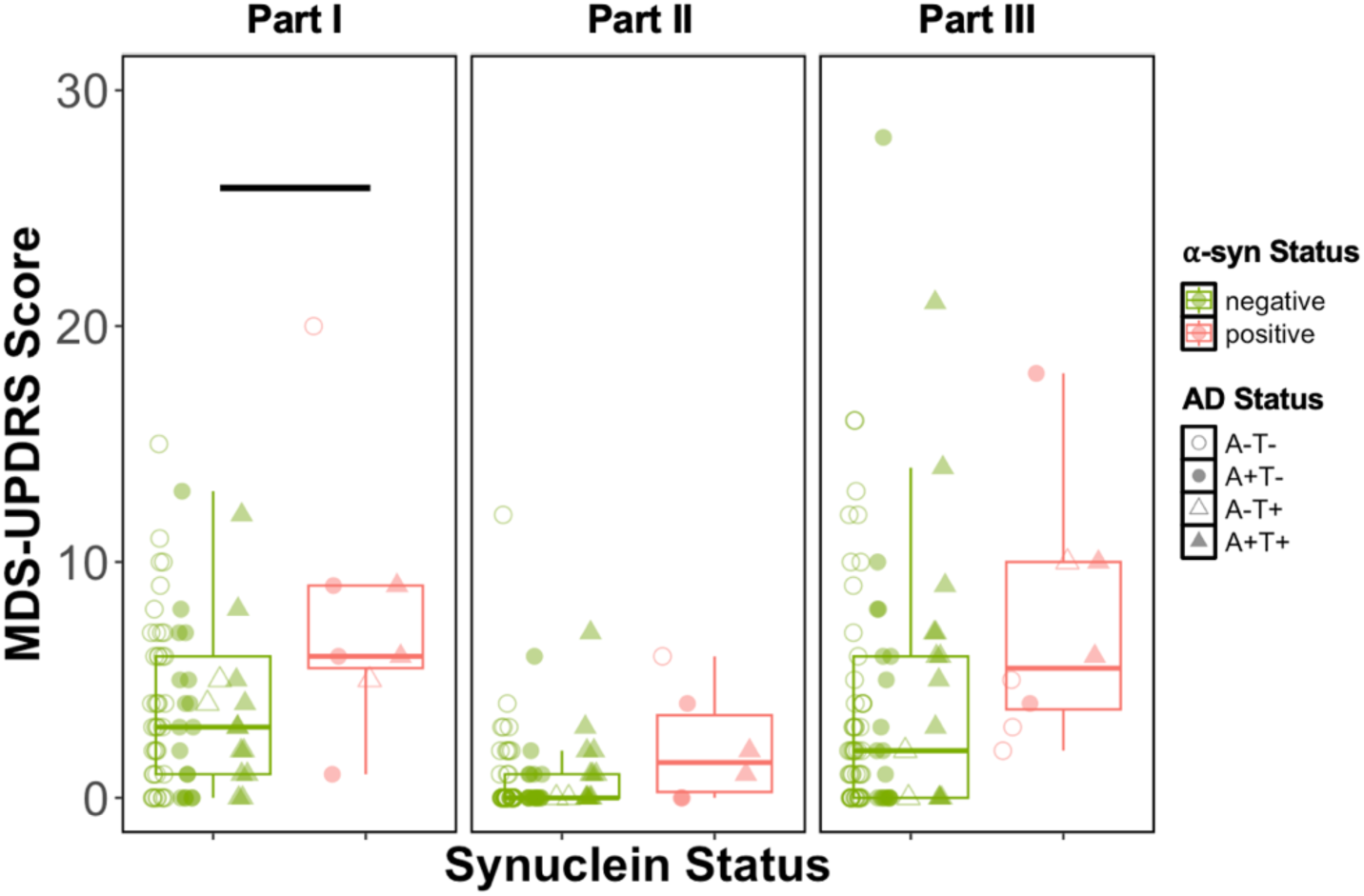
α-synuclein associations with Parkinson’s disease symptom severity in clinically unimpaired participants. Boxplots show median and interquartile range in Movement Disorders Society Unified Parkinson’s Disease Rating Scale (MDS-UPDRS) (A) Part I non-motor symptoms, (B) Part II self-reported motor symptoms, and (C) Part III motor exam for α-synuclein positive and negative clinically unimpaired participants. Unadjusted data are displayed. Solid line indicates significant difference in a linear model adjusting for age and sex. One outlier is omitted from the Part III plot, a α-synuclein negative participant with a score of 43.

Finally, in a subset of CU who had neuropsychiatric symptom severity assessed with the NPI-Q, α-syn positivity was not associated with NPI-Q severity in a linear regression that included age and sex (0.07 ± 0.25, p=0.77). Similarly, there was no association with NPI-Q severity score when individuals with AD-MCI and AD-Dementia were included in the analysis with diagnosis as an additional covariate (0.15 ± 0.17, p=0.38).

## Discussion

We found that 9% of CU, 14% of AD-MCI, and 19% of AD-Dementia participants were α-syn SAA positive, versus 81% of individuals with LBD spectrum clinical diagnoses. We found that α-syn+ CU older adults were older, performed worse on tests of executive function and working memory, and reported more non-motor symptoms associated with LBD relative to α-syn-CU adults. In contrast, α-syn status was not significantly associated with CSF Aβ or tau, memory performance, motor symptoms, or more severe neuropsychiatric symptoms. These cross-sectional findings represent an important addition to the limited but growing literature characterizing the frequency and effects of α-syn positivity in CU older adults and in individuals with AD who are assessed with CSF α-syn SAA.

The frequency of α-synuclein positivity in individuals above age 50 is estimated to be 8-14% based on postmortem neuropathology studies, which includes those with brainstem only Lewy body pathology and not necessarily cortical pathology^13,14^. Given this low prevalence, large samples are necessary to identify sufficient asymptomatic α-syn+ CU individuals to detect subtle effects of preclinical pathology. Furthermore, detection of *in vivo* α-synuclein through SAA is currently only possible with CSF (in clinical practice), which is only available in select healthy aging cohorts. To date, the BioFINDER study is the largest of α-syn SAA in CU older adults, which found a frequency of 8% α-syn+ in N=1,182 older adults with a mean age of 70^10^. Beyond BioFINDER, 16% of N=576 CU (mean age 74) in the Alzheimer’s Disease Neuroimaging Initiative (ADNI) were α-syn+^11^, and 9% of N=378 CU (mean age 65) were α-syn+ in aging cohorts from University of Wisconsin^12^ (also see smaller studies^30–32)^. The ADNI CU cohort was older and had higher frequency of Aβ positivity relative to the present Stanford cohort, BioFINDER, and the Wisconsin cohort, though it is not entirely clear whether these factors might explain why the rate of α-syn positivity is twice as high in ADNI. Regardless of possible differences across cohorts, it is notable that the 8-16% CU α-syn positivity frequency assessed with CSF α-syn SAA in these four cohorts is similar to the 8-14% frequency reported in the postmortem literature^13,14^. Collectively, these data support inclusion of α-syn SAA positive individuals, who do not yet manifest neurological symptoms, within an integrated staging system for NSD (NSD-ISS, stage 1)^3^. This prodromal group represents a key opportunity to understand the effects of initial pathological processes relevant for LBD and provides opportunities for future prevention strategies.

We found that α-syn+ CU were older compared to α-syn-CU, replicating previous CSF α-syn SAA findings^10–12^. In our study, males were not significantly more likely to be α-syn+ (OR 1.42, 95% CI 0.59-3.43, adjusted for age). This lack of a sex difference is in contrast with the BioFINDER study, which found males were more than twice as likely to be α-syn+ than females (OR 2.56, adjusted for age)^10^, as well as the Wisconsin study^12^, although significant sex differences were not present in ADNI^11^. In terms of sex differences in rates of clinical LBD, the incidence of DLB is higher in males^33^ and it has been estimated that the prevalence of PD is as much as 1.5-2 times higher in males. However, a meta-analysis of 32 studies noted that this gap differs across countries and may be shrinking^34^. In participants with AD, the α-syn+ rates of 14% in MCI and 19% in dementia observed our cohort were lower than in BioFINDER (19% MCI, 28% dementia)^35^ and ADNI (19% MCI, 38% dementia)^11^, potentially due to participants at Stanford being evaluated for signs and symptoms indicative of clinical LBD. While we observed α-syn positivity to be more frequent with increasing clinical impairment (CU versus AD-MCI and Dementia-AD), this was not statistically significant in an adjusted logistic regression, possibly due to low sample size in the AD subsamples. Finally, there was no significant difference in years of education in CU α-syn+ versus α-syn-status in our study, which was also the case in the Wisconsin cohort and ADNI^11,12^ and not reported in BioFINDER^10^.

Postmortem studies indicate that α-synuclein and AD pathology frequently co-occur, both in individuals with clinical diagnoses of AD^36,37^ and LBD^38,39^, and rodent studies suggest that Aβ may promote the spread of α-synuclein along with tau^40^. We observed that a greater percentage of Aβ+ CU were α-syn+ relative to Aβ-CU (13% vs. 7%), though this difference was not statistically significant. However, our α-syn+ odds ratio of 1.81 in Aβ+ versus Aβ-CU is comparable to the effect observed in the larger BioFINDER (OR 1.72, adjusting for age) and ADNI studies (OR 2.03)^10,11^. The effect for tau positivity predicting α-syn SAA status also was not significant (OR 1.40). Tau positivity was not associated with α-syn+ in BioFINDER CU^10^, although the Wisconsin study observed an association (n.b., 30 of the 411 participants in that analysis had MCI, which may have influenced the association)^12^. Interestingly, in ADNI the effect of tau positivity was observed in the opposite direction, with tau+ CU less likely to be α-syn+^11^. When considering continuous measures of Aβ and tau (rather than dichotomous status) in CU, we similarly did not observe significant differences by α-syn SAA status. A study combining data from 2,315 participants across BioFINDER and ADNI found that α-syn positivity was associated with higher Aβ PET and lower CSF Aβ, suggesting greater Aβ aggregation, but found no association with tau^41^. In a different approach to the ADNI data, another group found that Aβ+/α-syn+ participants spanning the full AD clinical continuum had higher levels of tau and accumulated tau faster over a 2.5-year period relative to Aβ+/α-syn-participants, suggesting α-synuclein co-pathology contributes to AD progression^42^. In summary, these findings support the hypothesis that α-synuclein and Aβ pathology may co-occur even before the onset of symptoms. The incongruent findings with tau positivity in CU between the BioFINDER, Wisconsin, and ADNI cohorts suggest that in vivo mixed-pathology effects are complex and may depend on the inclusion of later clinical stages (ie. MCI, dementia) and/or other cohort differences. Collectively, these findings support the efforts to include biomarkers of additional pathologies when considering a biological definition of neurodegenerative diseases, as now included in the revised criteria for diagnosis and staging of AD^9^ and highlighted as an outstanding research question that warranted further investigation in the NSD-ISS^3^.

APOE ε4 is the strongest genetic risk factor for sporadic AD. Postmortem studies suggest that APOE ε4 carriers with LBD are more likely to have AD co-pathology^43^ but not more likely to have LBD in isolation^44^. We found that when adjusting for age, CU APOE ε4 carriers were more than twice as likely to be α-syn+, although this effect was attenuated when adjusting for Aβ status. The Wisconsin study did not observe differences in α-syn SAA positivity when comparing APOE genotype^12^, and the ADNI study showed an increased frequency of α-syn+ only in APOE ε4/ε4 homozygotes, which was attenuated when adjusting for Aβ^11^. In our study, 48% APOE ε4/ε4 homozygotes were α-syn+. BioFINDER did not report APOE status in their analyses^10^. Future studies should investigate the interactions between *APOE* ε4, α-syn, and AD across aging and cognitive decline.

Interestingly, we found that α-syn+ CU performed worse than α-syn-CU on tests of executive function and working memory even after adjusting for continuous CSF p-tau181. In contrast, p-tau181 was associated with worse episodic memory performance. There are limited postmortem data investigating the cognitive effects of α-synuclein pathology in CU older adults. In contrast to the memory changes typically observed in AD, executive function and visuospatial domains are more likely to be affected in clinical LBD^20,45^. Notably one small study of 15 individuals with autopsy-confirmed Lewy body pathology, but without clinical impairment at death, found abnormal performance on Trails B, suggesting executive function impairment^15^. In the BioFINDER study, α-syn+ CU had worse cross-sectional performance in global cognition as well as episodic memory performance and showed greater longitudinal decline in these domains in addition to attention/executive function^10^. These patterns were similar when comparing individuals with and without AD pathology, suggesting a contribution of Lewy body pathology independent of AD. In ADNI, there were no cross-sectional or longitudinal differences in CU cognitive performance by α-syn SAA status^11^. Our study was limited by the number of cognitive tests available across all participants and did not have a test of visuospatial function, which is commonly affected by Lewy body pathology^20,45^. More work is needed to understand which cognitive domains are most sensitive to α-synuclein pathology in the presymptomatic stage.

α-syn+ CU participants endorsed significantly higher non-motor symptom severity on the MDS-UPDRS Part I relative to α-syn-CU, suggesting that α-syn positivity increases the likelihood of non-motor symptoms in the preclinical phase. Part I of the UPDRS captures mood symptoms including anxiety, depression, apathy, in addition to other LBD-related symptoms including constipation, fatigue, and sleep dysfunction. Sense of smell reduced in α-syn+ CU in the BioFINDER study^10^ and smell dysfunction is a robust predictor of α-syn positivity in clinical LBD cohorts^46,47^. Unfortunately, we did not have a measure of smell function in our cohort. We did not observe a significant difference by α-syn SAA status in self-reported or neurologist-assessed motor function (MDS-UPDRS parts II & III) in CU, which replicates similar findings in BioFINDER^10^. In contrast, a study of hyposmic older adults without a diagnosis of PD found that α-syn positivity was associated with worse MDS-UPDRS motor function (part III)^16^. When including participants with AD in the analysis, there were associations between α-syn positivity and non-motor symptoms as well as self-reported motor symptoms. Similarly, motor function was more impaired in α-syn+ individuals with AD in BioFINDER^35^. We hypothesized that α-syn+ individuals would have higher neuropsychiatric symptom severity, however there was no α-syn-related difference in scores on the NPI-Q in CU or AD. Tosun and colleagues similarly reported no differences on the hallucinations and sleep disturbances items of the NPI-Q in ADNI on the basis of α-syn SAA status in CU or AD^11^, although individuals with AD in BioFINDER who were α-syn+ were more likely to endorse experiencing hallucinations^35^.

One consideration in interpreting these findings is the sensitivity of CSF α-syn SAA, which has been validated against postmortem results in a limited number of studies ^6–8,11,48^. Postmortem comparison studies consistently show that α-syn SAA has high sensitivity (∼100%) in detecting cortical Lewy bodies but only around 50% sensitivity in detecting isolated brainstem and amygdala-only Lewy body pathology (with the exception of individuals with REM-sleep behavior disorder^49^). The implication for examining SAA α-syn+ CU individuals is that we may be missing as many as 50% of individuals in the early (pre-cortical) pathological stages of LBD and therefore underestimating the prevalence of α-syn positivity, and thus NSD-ISS stage 1, in the broader population.

An important limitation of our study was that it was cross-sectional, and we did not have follow up data to determine whether individuals developed clinically specific LBD symptoms or clinically declined over time. Previous studies indicate that α-syn+ CU are likely to progress to PD or DLB^10^ and that α-syn+ AD are more likely to develop LBD signs and symptoms^50^. While existing cohorts with CSF α-syn SAA data tend to focus either on the AD spectrum or LBD spectrum, future work could aim to bridge clinical categories to understand the effects of α-syn positivity agnostic to current clinical diagnostic frameworks. In identifying demographic, cognitive, and clinical outcomes associated with α-syn positivity in unimpaired older adults, our findings add converging evidence for including individuals with the earliest signs of α-synuclein deposition in a biological framework.

## Data Availability

Anonymized data will be made available on request to qualified researchers who have Stanford University's institutional review board approval and a Data Usage Agreement.

## Study Funding

This work was supported by the National Institutes of Health under award numbers P50AG047366, P30AG066515, R01AG048076, R01AG074339, K99AG071837, and the National Centralized Repository for Alzheimer’s Disease and Related Dementias (NCRAD) grant U24AG021886. Support was also provided by The Phil and Penny Knight Initiative for Brain Resilience, The Sue Berghoff LBD Research Fellowship, and the Alzheimer’s Association under award number AARFD-21-849349.

## Disclosure

MJP is currently a full-time employee at Amprion Inc.

## Table

|  | **Clinically unimpaired** |  | **Alzheimer's disease** |  |
| --- | --- | --- | --- | --- |
| **α-synuclein SAA status** | **Negative** | **Positive** | **Negative** | **Positive** |
| **N (%)** | 246 (91.1%) | 24 (8.8%) | 47 (83.9%) | 9 (16.1%) |
| **Age (years)** | 68.3 (7.07) | 72.3 (6.82) | 67.4 (8.55) | 69.0 (8.47) |
| **N (%) Female** | 146 (59.3%) | 11 (45.8%) | 24 (50.0%) | 3 (33.3%) |
| **Years of education** | 16.9 (2.19) | 16.8 (2.50) | 16.1 (2.36) | 17.4 (2.88) |
| Missing | 26 (10.6%) | 1 (4.2%) | 1 (2.1%) | 1 (11.1%) |
| **N (%) Impairment status** |  |  |  |  |
| Normal | 246 (100%) | 24 (100%) | 0 (0%) | 0 (0%) |
| MCI | 0 (0%) | 0 (0%) | 25 (53.2%) | 4 (44.4%) |
| Dementia | 0 (0%) | 0 (0%) | 22 (46.8%) | 5 (55.6%) |
| **N (%) AD status** |  |  |  |  |
| A-T- | 144 (58.5%) | 11 (45.8%) | 0 (0%) | 0 (0%) |
| A+T- | 47 (19.1%) | 7 (29.2%) | 11 (22.9%) | 1 (11.1%) |
| A-T+ | 8 (3.3%) | 1 (4.2%) | 0 (0%) | 0 (0%) |
| A+T+ | 30 (12.2%) | 4 (16.7%) | 37 (77.1%) | 8 (88.9%) |
| Missing | 17 (6.9%) | 1 (4.2%) | 0 (%) | 0 (%) |
| **N (%) APOE genotype** |  |  |  |  |
| ε2/ε3 | 22 (8.9%) | 1 (4.2%) | 3 (6.3%) | 0 (0%) |
| ε2/ε4 | 4 (1.6%) | 0 (0%) | 1 (2.1%) | 0 (0%) |
| ε3/ε3 | 122 (49.6%) | 9 (37.5%) | 7 (14.6%) | 2 (22.2%) |
| ε3/ε4 | 44 (17.9%) | 5 (20.8%) | 24 (50.0%) | 1 (11.1%) |
| ε4/ε4 | 7 (2.8%) | 4 (16.7%) | 3 (6.3%) | 4 (44.4%) |
| Missing | 47 (19.1%) | 5 (20.8%) | 10 (20.8%) | 2 (22.2%) |

Participant characteristics based on clinical group and α-synuclein status. Mean and standard deviation shown for age and years of education. Alzheimer's disease status assessed with CSF β-amyloid 42/40 (cutoff <0.0943) and p-tau181 (cutoff >57.5 pg/mL). MCI, mild cognitive impairment; AD, Alzheimer's disease; A/T, β-amyloid/tau; SAA, seed amplification assay.

## Notes

### Author Declarations

The Institutional Review Board of Stanford University gave ethical approval for this work.

