## Supplemental File for "Effects of α-synuclein pathology in normal aging and Alzheimer’s disease"

**Supplementary Methods**

**CSF ⍺-synuclein SAA batch overview**

Overall, 472 CSF samples from 426 unique participants were sent to Amprion, Inc. After removing longitudinal data and applying exclusion criteria based on clinical characteristics, CSF a-syn SAA results from 416 total participants were included in the present cross-sectional analyses.

Batch 1: 154 samples from 153 unique participants were processed by Amprion, Inc in June 2022. 151 of these participants were included in a previous publication^1^. In this initial analysis, we identified 17 participants with unexpected clinical-biomarker profiles^1^ and re-ran the SAA (Batch 2) on a separate CSF aliquot collected during the same lumbar puncture. Three additional participant CSF samples were also processed in both batches due to study co-enrollment, for a total of 20 samples repeated across batches. Of these 20 cases assayed in both Batch 1 and Batch 2, 5 switched CSF α-syn status. We suspect status change was driven by the fact that these re-assayed CSF samples were intentional selected given their clinicobiomarker discrepancy and does not reflect the overall test-retest reliability of the a-syn SAA. Because repeat testing was not conducted across the entire cohort, ⍺-syn status for these 5 participants was defined as their Batch 1 result for analyses in the current manuscript. One MCI-AD participant processed in Batch 1 had a longitudinal CSF sample also assayed in Batch 1 (delay between CSF samples = 1.0 years); both results were CSF ⍺-syn-.

Batch 2: 318 CSF samples from 296 unique participants were processed by Amprion, Inc in January 2024. Two PD participants with baseline CSF samples in Batch 1 had longitudinal CSF samples in Batch 2 due to co-enrollment between PUC and ADRC (mean delay between samples=3.8 ± 0.3 years). These participants were CSF ⍺-syn+ at both time points. One CU participant CSF sample was included twice in Batch 2 due to co-enrollment in ADRC and SAMS; both results were CSF ⍺-syn-. One MCI-AD participant had a baseline and longitudinal CSF sample assayed in Batch 2 (delay between samples = 2.9 years); both results were CSF ⍺-syn-. Seven CU participants from SAMS had 2 longitudinal CSF samples (mean delay between samples = 5.3 ± 2.1 years) and six had 3 longitudinal CSF samples (mean delay between first and last samples = 7.3 ± 0.2 years). One CU SAMS participant had a baseline CSF sample processed in batch 1 and two longitudinal CSF samples processed in batch 2 (delay between first and last samples = 4.3 years). Of these 15 CU participants with longitudinal assessments, 13 were CSF α-syn- across all time points, and 1 was CSF α-syn- during their first visit and CSF α-syn+ 3.4 years later; for this participant all demographic and clinical data were obtained from the SAMS time point corresponding to the CSF α-syn+ result (Wave 1.5), and the individual was considered CSF α-syn+ in all analyses. One participant with a longitudinal CSF sample was indeterminate at their first visit and was CSF α-syn- 7.6 years later; this participant was classified as CSF α-syn- in our analyses. 10 participants with clinically determined AD diagnoses (MCI-AD or Dementia-AD) were excluded from analyses because they did not have evidence of CSF amyloid positivity (3 CSF Aβ-, 7 missing CSF Aβ42/40). After removing these 10 AD participants, the 20 CSF samples repeated across batches 1 and 2, 1 CSF sample processed twice, and 24 longitudinal CSF samples, there were a total of 263 participants processed in Batch 2 included in the manuscript.

**Supplementary Tables**

**Supplementary Table 1 – Participants with consensus clinical diagnoses other than clinically unimpaired, Alzheimer’s disease, or Lewy body disease.**

| **Clinical diagnosis** | **Biomarker status** |
| --- | --- |
| Frontotemporal dementia | ⍺-syn-, A+T- |
| Mild cognitive impairment due to depression | ⍺-syn-, A+T- |
| Mild cognitive impairment due to cardiovascular disease | ⍺-syn-, A-T- |
| Mild cognitive impairment, not otherwise specified | ⍺-syn-, A+T+ |
| Mild cognitive impairment, not otherwise specified | ⍺-syn+, A-T- |

All five participants were enrolled in the Stanford Alzheimer’s Disease Research Center.

**Supplementary Table 2 – Lewy body disease participant demographics.**

|  | **Cognitively unimpaired** | | **Mild cognitive impairment** | | **Dementia** | |
| --- | --- | --- | --- | --- | --- | --- |
| **CSF ⍺-synuclein status** | **Negative** | **Positive** | **Negative** | **Positive** | **Negative** | **Positive** |
| **N (%)** | 6 (18.8%) | 32 (81.2%) | 7 (23.3%) | 23 (76.7%) | 3 (17.6%) | 14 (82.3%) |
| **Age (years)** | 68.3 (6.02) | 66.2 (7.49) | 72.4 (9.69) | 69.6 (5.79) | 74.3 (14.0) | 71.1 (6.78) |
| **N (%) Female** | 2 (33.3%) | 17 (53.1%) | 3 (42.9%) | 7 (30.4%) | 2 (66.7%) | 3 (21.4%) |
| **Years of education** | 16.8 (2.23) | 16.6 (2.27) | 18.7 (1.25) | 16.3 (2.72) | 15.3 (3.06) | 16.5 (2.18) |
| **N (%) Clinical diagnosis** |  |  |  |  |  |  |
| Parkinson’s disease | 6 (100%) | 32 (100%) | 5 (71.4%) | 20 (87.0%) | 1 (33.3%) | 10 (71.4%) |
| Dementia with Lewy bodies | 0 (0%) | 0 (0%) | 2 (28.6%) | 3 (13.0%) | 2 (33.3%) | 4 (28.5%) |
| **N (%) AD status** |  |  |  |  |  |  |
| A-T- | 5 (83.3%) | 25 (78.1%) | 4 (57.1%) | 9 (39.1%) | 0 (0%) | 3 (21.4%) |
| A+T- | 1 (16.7%) | 5 (15.6%) | 0 (0%) | 8 (34.8%) | 0 (0%) | 4 (28.6%) |
| A-T+ | 0 (0%) | 0 (0%) | 1 (14.3%) | 0 (0%) | 0 (0%) | 0 (0%) |
| A+T+ | 0 (0%) | 1 (3.1%) | 2 (28.6%) | 5 (21.7%) | 3 (100%) | 6 (42.9%) |
| Missing | 0 (0%) | 1 (3.1%) | 0 (0%) | 1 (4.3%) | 0 (0%) | 0 (0%) |
| **N (%) APOE genotype** |  |  |  |  |  |  |
| ε2/ε3 | 1 (16.7%) | 1 (3.1%) | 2 (28.6%) | 3 (13.0%) | 1 (33.3%) | 1 (7.1%) |
| ε2/ε4 | 0 (0%) | 1 (3.1%) | 0 (0%) | 0 (0%) | 0 (0%) | 0 (0%) |
| ε3/ε3 | 3 (50.0%) | 17 (53.1%) | 5 (71.4%) | 14 (60.9%) | 1 (33.3%) | 5 (35.7%) |
| ε3/ε4 | 1 (16.7%) | 9 (28.1%) | 0 (0%) | 5 (21.7%) | 0 (0%) | 5 (35.7%) |
| ε4/ε4 | 1 (16.7%) | 0 (0%) | 0 (0%) | 1 (4.3%) | 0 (0%) | 1 (7.1%) |
| Missing | 0 (0%) | 4 (12.5%) | 0 (0%) | 0 (0%) | 1 (33.3%) | 2 (14.3%) |

Characteristics for participants with Lewy body disease diagnoses based on cognitive impairment severity and CSF ⍺-synuclein status. Alzheimer’s disease status assessed with CSF amyloid 42/40 and p-tau181. MCI, mild cognitive impairment; AD, Alzheimer’s disease; A/T, amyloid/tau.

**Supplementary Table 3 – Neuropsychological test performance and associations with a-synuclein SAA and p-tau181 in clinically unimpaired participants.**

| **Neuropsychological Test** | **Mean (SD)** | | **a-synuclein SAA**  **β ± SE, p-value** | **P-tau181**  **β ± SE, p-value** |
| --- | --- | --- | --- | --- |
|  | **a-synuclein SAA negative** | **a-synuclein SAA positive** |  |  |
| **Digit Span Forward**^2,3^ | 10.0 (2.33) | 9.71 (2.59) | 0.07 ± 0.54, 0.89 | 0.00 ± 0.01, ± 0.73 |
| **Digit Span Backward**^2,3^ | 7.75 (2.24) | 6.52 (2.23) | -0.97 ± 0.52, 0.07 | -0.01 ± 0.01, 0.14 |
| **Trail Making Test A (s)**^4^ | 30.8 (9.26) | 30.6 (5.17) | -1.00 ± 1.97, 0.61 | 0.05 ± 0.02, 0.04 |
| **Trail Making Test B-A (s)**^4^ | 41.4 (24.2) | 56.0 (34.9) | 11.2 ± 4.95, 0.02 | 0.03 ± 0.06, 0.58 |
| **HVLT-R Delayed Recall**^5^ | 9.88 (2.32) | 8.81 (3.22) | -0.68 ± 0.54, 0.21 | -0.02 ± 0.02, 0.001 |
| **Semantic Fluency**^6^ | 23.9 (6.05) | 23.3 (5.89) | -0.07 ± 1.35, 0.96 | -0.01 ± 0.02, 0.41 |

Mean (SD) values for each group are not adjusted. Linear regression models included age, sex, and years of education as covariates. Unstandardized estimates and standard error (SE) are shown.
